# Mycobacterium tuberculosis whole genome sequence data support repurposing antileprosy antibiotic as antituberculosis

**DOI:** 10.1101/2020.08.15.20175570

**Authors:** Jamal Saad, Jenny Gallou, Nathalie Brieu, Michel Drancourt, Sophie Alexandra Baron

**Author notes:** **Corresponding author**: Sophie Alexandra Baron, Aix Marseille Univ, IRD, APHM, MEPHI, IHU Méditerranée Infection, Faculté de Médecine et de Pharmacie, 19-21 boulevard Jean Moulin, 13385 Marseille CEDEX 05, France.

## Abstract

**Background:** We implanted WGS as the routine method to profile the antibiotic susceptibility of *M. tuberculosis* isolates focusing on *in silico* resistance to antileprosy drugs that we recently proposed to reposition for the treatment of pulmonary tuberculosis.

**Methods:** We prospectively performed WGS of 112 *M. tuberculosis* isolates recovered from respiratory tract samples of 106 patients diagnosed with pulmonary tuberculosis between 2017 and 2019 and defined their antibiotic susceptibility profile to 17 antibiotics including antileprotics drugs.

**Results:** We incidentally observed 08 sequence variations in 07 genes, specific to seven sublineages. Altogether, we observed 09 (8%) rifampicin-resistant, 05 (4.4%) multidrug-resistant and 02 (1.7%) extensively-drug resistant isolates; whereas only one isolate exhibited in silico resistance to clofazimine.

**Conclusion:** These results support repurposing of antileprosis antibiotics as antituberculosis; and offer new targets for genotyping *M. tuberculosis*.

## Introduction

Point-of-care molecular assays detecting *Mycobacterium tuberculosis* DNA directly in respiratory tract samples, sharply decreased delay for pulmonary tuberculosis diagnosis (1). Some assays are also detecting the resistance of *M. tuberculosis* to rifampicin, the cornerstone drug for combined treatment of pulmonary tuberculosis (2). Further antituberculosis antibiotic susceptibility profiling currently relies upon standardized antituberculosis susceptibility tests, apart from the line probe assays detecting potential resistance to rifampicin, isoniazid, fluoroquinolones and aminoglycosides (3,4). GenXpert® (Cepheid Inc., Sunnyvale, CA, USA) and line probe assays however explore a limited number of genome sequence loci, complicating the interpretation of these assays and potentially leading to inaccurate positive results (3–6). Also, mixed *M. tuberculosis* strains may result in false-negative detection of rifampin resistance using the GenXpert® assay (7). Whole genome sequencing (WGS) entered the mycobacteriology laboratory to provide accurate identification and genotyping of *M. tuberculosis* isolates (8). WGS data have been secured in large databases for epidemiology purposes (9–12) and recent study indicated that WGS information could also be used to derive antibiotic susceptibility and resistance profile, eventually surrogating AST (13). Accordingly, we implanted WGS as the routine method to profile the antibiotic susceptibility of *M. tuberculosis* isolates and we observed in our local collection of *M. tuberculos*is isolates, the rarity of *in silico* resistance to antileprosy drugs that we recently proposed to reposition for the treatment of pulmonary tuberculosis (14).

## MATERIALS AND METHODS

### Ethic Statement

This study was qualified as a retrospective study that used not involving the human being. According to French ethics law, retrospective study did not require approval from a Research Ethics Committee, but they must be declared or covered by reference methodology of the National Institute of Health Data (“Institut National des Données de Santé”, INDS). After evaluation and validation by the data protection officer and according to the General Data Protection Regulation, this study, completing all the criteria, has been registered in the register of Marseille University Hospital (AP-HM) and cover by the MR-004 (INDS n° MR 0212080720).

### Clinical isolates

A total of 112 clinical *M. tuberculosis* isolates have been recovered from 106 patients diagnosed with pulmonary tuberculosis and 10 patients diagnosed with extra-pulmonary tuberculosis including 02 disseminated tuberculosis with pulmonary and lymph node, 01 tuberculous meningitis and 03 lymph node tuberculosis. These isolates have been cultured in the Mycobacteriology Reference Laboratory of the IHU Méditerranée Infection, Marseille, France in 2017 (22 isolates), 2018 (51 isolates) and 2019 (39 isolates).

### Genome sequencing and bioinformatical analysis

Isolates were subcultured on Middlebrook 7H10 agar medium supplemented with 10% oleic acid-albumin-dextrose-catalase (Becton Dickinson, Sparks, USA). Total DNA extracted by using InstaGen matrix (BioRad, Marnes-la-Coquette, France) and sequenced by Illumina MiSeq runs using a paired-end strategy (Illumina Inc., San Diego, USA) as previously described (15). The genetic support for altered susceptibility was examined for the following drugs: amikacin (AMK); bedaquiline (BDQ); capreomycin (CAP); chloramphenicol (CHL); clofazimine (CLO); ethambutol (EMB); ethionamide (ETO); fluoroquinolones (FLQs); isoniazid (INH); kanamycin (KAN); linezolid (LZD); para-aminosalicylic acid (PAS); pyrazinamide (PZA); rifampicin (RIF); and streptomycin (STR). Antibiotic resistance was detected directly on output MiSeq reads using three databases including Tb-profiler (10,11), TGS-TB (Total Genotyping Solution for Mycobacterium tuberculosis ver. 2) and Mykrobe Predictor TB_v0.1.3 (9,16).

## RESULTS AND DISCUSSION

### Presence of genotype-specific mutations

The 112 *M. tuberculosis* isolates were scattered among 04 lineages and 25 sublineages of *M. tuberculosis* (Supplementary Table S1), thus offering a reasonable diversity compared to what is currently described in this species (11). Unanticipatedly, the investigation of the 112 WGS yielded previously unreported mutations and indels that were seemingly specific to seven lineages/sublineages. In details, we observed a silent mutation C195T in the *pnc*A gene (pyrazinamide) in 08 isolates all belonging to sublineage L3.1.1 (East-African-Indian); a C391T/131-Q deletion in the *sir*R-Rv2788 gene (chloramphenicol) detected in 03 isolates all belonging to lineage L1 (L1, L1.1, L1.1.2) (Table S1); a C198T mutation in Rv0191 gene (chloramphenicol) detected in 30 isolates specific to sublineage L4.1.2 (3 isolates) and L4.1.2.1 (27 isolates) and a G139C deleterious mutation in Rv1353c gene (chloramphenicol) detected in 04 isolates of the sublineage L4.3.3 isolates; a mutation A857G/ D286G and insertion R473_V474insR in Rv17979c gene (bedaquiline-clofazimine) specifically detected in lineage L1 isolates (L1, L1.1, L1.1.2) (Indo-Oceanic) and in 05 isolates sublineage L4.6.2.2 (Euro-American genotype Cameroon), respectively. Finally regarding 14 isolates of the Beijing lineage (East-Asian, more precisely sublineage L2.2.1), we observed for the first time a A636C silent mutation in the *rps*A gene (pyrazinamide) and a G637A mutation in the in Rv0191 gene (chloramphenicol). Furthermore, a 580-C insertion in *tap*-Rv1258c gene (tetracycline antibiotics class/pyrazinamide) previously reported in the Beijing family, was here specified as specific for the L2.2.1 sublineage of this Beijing family (17).

### Antibiotic susceptibility profile

The susceptibility profile of 12 antibiotics using the three available databases found 33/112 (29.4%) *M. tuberculosis* isolates exhibiting at least one predicted antibiotic resistance. In details, 18/112 (16%) isolates were predicted to be resistant to streptomycin, 17/112 (14.1%) to be resistant to isoniazid, 9/112 (8%) to be resistant to rifampicin and no detected isolates resistant to linezolid (LZD) and para-aminosalicylic acid (PAS) (Figure). We identified 05 multidrugresistant isolates, 02 extended-resistant isolates and no totally resistant isolate.

### Focus on antileprotics drugs susceptibility

After genome assembly using SPAdes 3.13.1 (18), we specifically investigated the susceptibility profile to major antileprotics; i.e clofazimine, minocycline and sulfonamide (19) along with pyrazinamide previously demonstrated to potentialize the mycobactericidal effect of clofazimine, *in vitro* and in animal models (Supplementary File) (20,21). Chloramphenicol and bedaquiline susceptibility profile were also analyzed. These genes were compared with homologous genes in the *M. tuberculosis* H37Rv (NC_000962.3). We also investigated tetracycline family resistance using CARD 2020 (22). The impact of the mutation on the protein function was estimated using the PROVEAN score (23); as detailed in the Supplementary data set (Supplementary File and Table S2). One isolate was predicted resistant to clofazimine but none to other antileprosy antibiotics sulfonamide and minocycline (24), or chloramphenicol; despite the fact that we observed mutations in resistant genes including some previously unreported lineage or sublineage-specific ones.

## CONCLUSION

WGS analysis in the routine clinical practice of any microbiology laboratory, in presence of suitable bioinformatics tools has the potential to detect resistance days before traditional phenotypic culture methods. Applying WGS directly to clinical samples could further reduce this delay for slowly growing pathogens, uncultured mycobacteria and fastidious ones (25–28). As protocols are emerging to allow WGS directly from respiratory tract samples, so that we anticipate that antibiotic susceptibility profiling of *M. tuberculosis* will be soon derivable directly from the clinical sample (25).

## Data Availability

The authors confirm that the data supporting the findings of this study are available within the article and its supplementary material

## Acknowledgment

We thank Gilles LIOTAUD, Ayse AKKAYA and technicians of our genomic platform for their technical support.

## Transparency declaration

The authors declare that they have no competing interests.

## Authors’ contributions

JS, MD and SAB designed the study and drafted and revised the manuscript.

JG, NB and performed microbiology analyses and collected clinical information.

JS and SAB performed *in silico* analyses.

All authors have read and approved the final manuscript.

## Tables and Figures

### Figure: 1

**Figure**. Heatmap view showing the profile of confirmed clinical *Mycobacterium tuberculosis* resistance strains (row min: no resistance, row max: resistance).

### Supplementary Data Set: 3

Supplementary File: Additional analysis of genes involved in resistance to pyrazinamide, clofazimine-bedaquiline, minocycline, sulfonamide and chloramphenicol.

Supplementary Table S1: List of lineages of which studied isolates belong.

Supplementary Table S2: Summary of genetic modifications observed in the genes analyzed and predicted function effect results (PROVEAN)

